# Age-stratified seroprevalence of antibodies against SARS-CoV-2 in the pre- and post-vaccination era, February 2020–March 2022, Japan

**DOI:** 10.1101/2022.07.11.22277481

**Authors:** Seiya Yamayoshi, Kiyoko Iwatsuki-Horimoto, Moe Okuda, Michiko Ujie, Atsuhiro Yasuhara, Jurika Murakami, Calvin Duong, Taiki Hamabata, Mutsumi Ito, Shiho Chiba, Ryo Kobayashi, Satoshi Takahashi, Keiko Mitamura, Masao Hagihara, Akimichi Shibata, Yoshifumi Uwamino, Naoki Hasegawa, Toshiaki Ebina, Akihiko Izumi, Hideaki Kato, Hideaki Nakajima, Norio Sugaya, Yuki Seki, Asef Iqbal, Isamu Kamimaki, Masahiko Yamazaki, Yoshihiro Kawaoka, Yuki Furuse

**Author notes:** Correspondence: Yamayoshi Seiya, DVM, PhD, Institute of Medical Science, University of Tokyo, 4-6-1 Shirokanedai, Minato-ku, Tokyo 108-8639, Japan, Yoshihiro Kawaoka, DVM, PhD, Research Institute, National Center for Global Health and Medicine, 1-21-1 Toyama, Shinjuku-ku, Tokyo 162-8655, Japan, Yuki Furuse, MD, PhD, Nagasaki University Graduate School of Biomedical Sciences, 1-12-4 Sakamoto, Nagasaki 852-8523, Japan.

## Abstract

Japan has reported a small number of COVID-19 cases relative to other countries. Because not all infected people receive diagnostic tests for COVID-19, the reported number of COVID-19 cases must be lower than the actual number of infections. Assessments of the presence of antibodies against the spike protein of SARS-CoV-2 can retrospectively determine the history of natural infection and vaccination. In this study, we assessed SARS-CoV-2 seroprevalence by analyzing over 60,000 samples collected in Japan from February 2020 to March 2022. The results showed that about 5% of the Japanese population had been infected with the virus by January 2021. The seroprevalence increased with the administration of vaccinations to adults; however, among the elderly, it was not as high as the vaccination rate, probably due to poor immune responses to the vaccines and waning immunity. The infection was spread during the epidemic waves caused by the SARS-CoV-2 Delta and Omicron variants among children who were not eligible for vaccination. Nevertheless, their seroprevalence was as low as 10% as of March 2022. Our study underscores the low incidence of SARS-CoV-2 infection in Japan and the effects of vaccination on immunity at the population level.

## Introduction

SARS-CoV-2, the etiological agent of COVID-19, emerged at the end of 2019 and caused a pandemic. As of April 2022, despite the development of effective vaccines, over 500 million people have been infected with the virus and about 6 million have died (1). In Japan, with a population of 125 million, the reported numbers are 7 million infections and 30,000 deaths (2); however, the actual number of infected people must be higher than the reported figure because not all infected people undergo diagnostic testing.

A serological survey can retrospectively find people who have been infected with the virus (3). Antibodies against the spike protein of SARS-CoV-2 are generated by vaccination and natural infection. In contrast, antibodies against other components of the virus, such as the nucleoprotein, represent a history of not vaccination with COVID-19 vaccines currently available in Japan but only natural infection with SARS-CoV-2. Analyses of seroprevalence in several countries have revealed that the actual incidence of SARS-CoV-2 infection is much higher than the reported COVID-19 cases (3). For example, in the US, the seroprevalence of antibodies against the nucleoprotein of SARS-CoV-2 ranged from 3% to 10% in 2020 (4–7), and this number reached roughly 20–60% in 2021 (8,9). However, diagnostic tests confirmed only a fraction of the infections, especially at the beginning of the pandemic. The ascertainment rate was < 1%–30% in 2020 (4–7) and increased to approximately 50% in 2021 (10).

Vaccines for SARS-CoV-2 can prevent severe illness and death from COVID-19 for those at high risk, such as the elderly (11). In addition, they can prevent viral infection and therefore have the potential to contribute to herd immunity and the containment of the disease (12). However, the continuous emergence of novel variants of SARS-CoV-2 and waning immunity have allowed the pandemic to linger (13,14).

Here, we report the seroprevalence of antibodies against the spike protein of SARS-CoV-2 obtained by analyzing over 60,000 samples from February 2020 to March 2022 in Japan. The results were compared to the number of reported COVID-19 cases to determine the actual incidence and the ascertainment rate. Furthermore, our findings reveal how vaccination influenced COVID-19 immunity in Japan at the population level.

## Methods

### Study subjects and samples

The study subjects were patients who visited Sapporo Medical University Hospital, Japanese Red Cross Ashikaga Hospital, Keio University Hospital, National Hospital Organization Saitama Hospital, Eiju General Hospital, Yokohama City University Medical Center, Yokohama City University Hospital, Keiyu Hospital, or Zama Children’s Clinic, Japan, between February 2020 and March 2022. Sapporo Medical University Hospital is located in Hokkaido Prefecture, whereas all of the other healthcare facilities are located in the Tokyo Metropolitan Area and its suburbs.

Residual serum or plasma samples collected for medical examination were analyzed for this study. The reason for the healthcare facility visit was not considered for inclusion in this study. However, patients positive for SARS-CoV-2 nucleic acid test or antigen test were excluded.

### Measurement of antibodies

An enzyme-linked immunosorbent assay (ELISA) to detect antibodies against SARS-CoV-2 was performed as described previously (15). Ninety-six-well Maxisorp microplates were incubated with 2 μg/ml of the recombinant receptor-binding domain (RBD) of the spike protein, the whole length of the nucleoprotein, or phosphate-buffered saline (PBS) at 4°C overnight. The microplates were then incubated with 5% skim milk in PBS containing 0.05% tween-20 (PBS-T).

The antigen-coated microplates were incubated with the serum or plasma samples 40-fold diluted in PBS-T, including 5% skim milk, followed by the peroxidase-conjugated goat anti-human IgG, Fcγ fragment specific antibody (Jackson ImmunoResearch Laboratories, West Grove, PA). One-step Ultra TMB-Blotting Solution (Thermo Fisher Scientific, Waltham, MA) was added to each well and incubated for 3 minutes at room temperature.

The reaction was stopped by adding 2M H_2_SO_4_, and the optical density at 450 nm (OD_450_) was immediately measured. The OD_450_ value of the PBS wells was subtracted from the OD_450_ value of the spike protein or nucleoprotein wells as background.

### Validation samples for ELISA

Convalescent sera from patients with laboratory-confirmed COVID-19 were used as positive controls, and residual serum samples collected in 2012 were used as negative controls to validate the ELISA tests.

### Other data sources

The daily number of reported COVID-19 cases was obtained from the website and press releases of each prefecture in the study area. Vaccine administration data were available in the Vaccination Record System (https://cio.go.jp/vrs). This system was launched in April 2021, and the number of vaccines administered before that time point was included on the first day of the record. All vaccines available in Japan require two doses for immunization; a third dose was administered as a booster after December 2021. The census data of Japan were downloaded and used to obtain demographic information in the study area (https://www.stat.go.jp/data/jinsui/2021np/index.html).

### Statistical analysis

A receiver operating characteristic curve (ROC) was drawn for the ELISA OD_450_ values to set the threshold to determine whether samples were negative or positive for SARS-CoV-2 spike protein and nucleoprotein by using Youden’s index (16).

The proportion of seropositive samples was investigated by month and age group. The Wilson 95% confidence interval was computed for the seroprevalence data (17). The numbers of reported COVID-19 cases and vaccine administrations per population were calculated by using the census data.

### Ethical considerations

The study protocol was reviewed and approved by the institutional review board of the Institute of Medical Science, University of Tokyo (protocol number 2019-75). The protocol was also checked and approved by each research institute and healthcare facility involved. The study participants gave informed consent during their healthcare facility visits for their data and residual samples to be used anonymously for clinical research.

## Results

During the study period, Japan had six COVID-19 epidemic waves (Figure 1). The cumulative number of confirmed COVID-19 cases by the end of March 2022 was 6.7 million in a population of 125 million. Vaccinations started in February 2021 for healthcare workers; then, in April 2021, they were expanded to the general population, prioritizing people at high risk, such as the elderly and those with comorbidities including respiratory disorders and immunocompromised diseases. A total of 256.8 million doses of vaccine were administered during the study period. The two mRNA vaccines, BNT162b2 and mRNA-1273, were the main vaccines administered in Japan.

**Figure 1.**
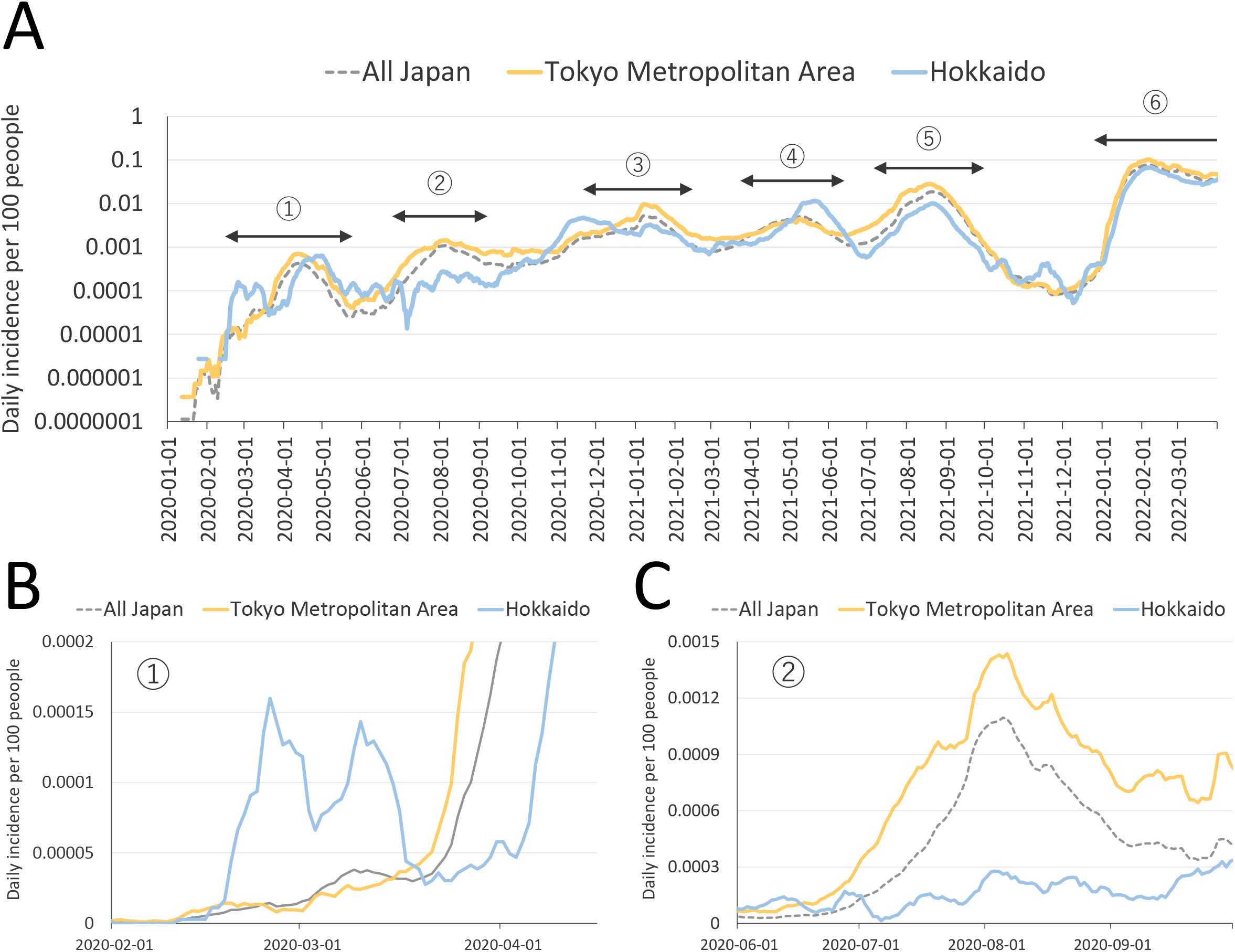
Epidemic curve of COVID-19 in Japan, January 2020–March 2022. A) The daily numbers of reported COVID-19 cases per 100 people in all of Japan, the Tokyo Metropolitan Area, and Hokkaido are shown. The numbers in the circles indicate the serial number of epidemic waves. The Y-axis is on a logarithmic scale. The fourth, fifth, and sixth waves were driven by the Alpha, Delta, and Omicron variants of SARS-CoV-2, respectively. B) and C) Parts of the first (B) and second (C) waves are depicted on a linear scale.

We first assessed SARS-CoV-2 antibody titers in prepandemic samples from 2012 and in samples of COVID-19 convalescent sera (Supplementary Figure 1). The thresholds for discriminating infected convalescent samples from uninfected prepandemic samples were determined by using ROCs. The specificity and sensitivity of the ELISA test for nucleoprotein antibodies were 98.0% and 95.6%, respectively.

The antibody titer for the RBD of the spike protein can be used to clearly differentiate convalescent samples from naïve samples, with a specificity of 99.5% and a sensitivity of 100%. Hence, we measured the antibody titers for the spike protein in our further analyses. It should be noted that our seroprevalence data cannot determine whether immunity was generated by natural infection or vaccination.

A total of 44,681 samples were collected in the Tokyo Metropolitan Area between February 2020 and March 2022. Of these samples, 44,672 (99.9%) were analyzed for the study, and 9 were excluded because the metadata were incomplete. The samples were collected from people aged 0– 105 years. The numbers of analyzed samples by age group and month are presented in Supplementary Table 1.

SARS-CoV-2 seroprevalence was low in 2020 in the Tokyo Metropolitan Area (Figure 2). In January 2021, just before the vaccine rollout, the proportions of serum samples positive for SARS-CoV-2 were 0%, 2.5%, 8.2%, 5.7%, 2.8%, 2.0%, 4.2%, 4.0%, and 3.7% in the 0–9, 10–19, 20–29, 30–39, 40–49, 50–59, 60–69, 70–79, and ≥ 80 age groups, respectively. The SARS-CoV-2 seroprevalence was significantly higher than the rate of cumulative reported COVID-19 cases in the population, as seen in points where the lower bound of the 95% confidence interval of the seroprevalence was higher than the cumulative incidence rate in Figure 2B.

**Figure 2.**
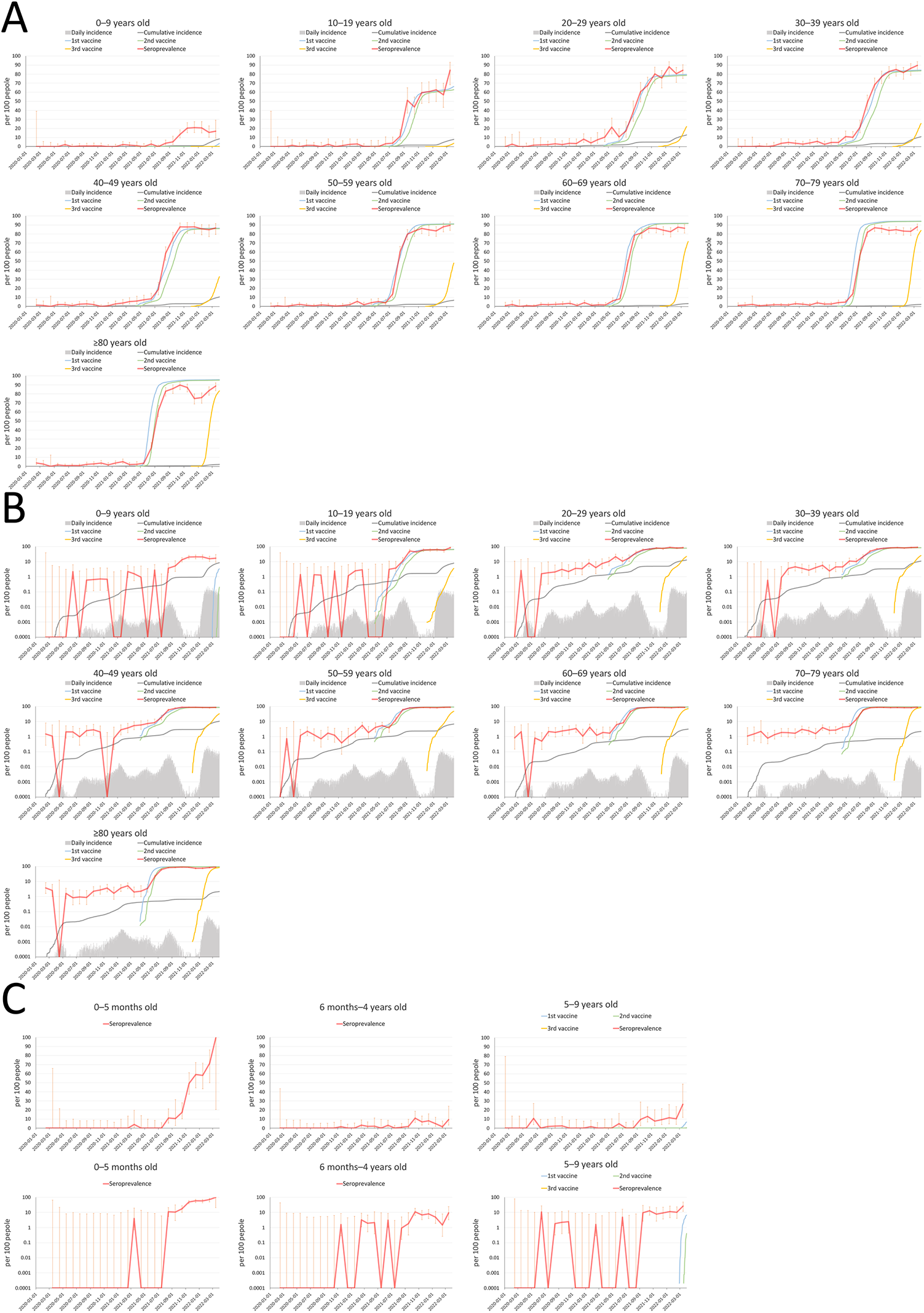
Seroprevalence of SARS-CoV-2 by age in the Tokyo Metropolitan Area, February 2020–March 2022. A) Seropositive rates for the SARS-CoV-2 spike protein in the Tokyo Metropolitan Area are shown by age group. The daily number of reported COVID-19 cases, their cumulative number, and the cumulative number for the first, second, and third vaccine administrations per population are also shown. The vertical orange lines indicate the 95% confidence intervals of the seroprevalence data. B) The same data on a logarithmic scale are shown. C) and D) The proportions of seropositive samples in children aged 0–6 months, 6 months–4 years, and 5–9 years are shown on linear (C) and logarithmic (D) scales.

We then calculated the ratio of the seroprevalence to the cumulative incidence by the time of vaccination for the general public (Figure 3). This rate can correspond to the number of actual infected persons per detected case. However, a low antibody titer in some infected persons due to a weak immune response and waning immunity can affect the accuracy of the estimation.

**Figure 3.**
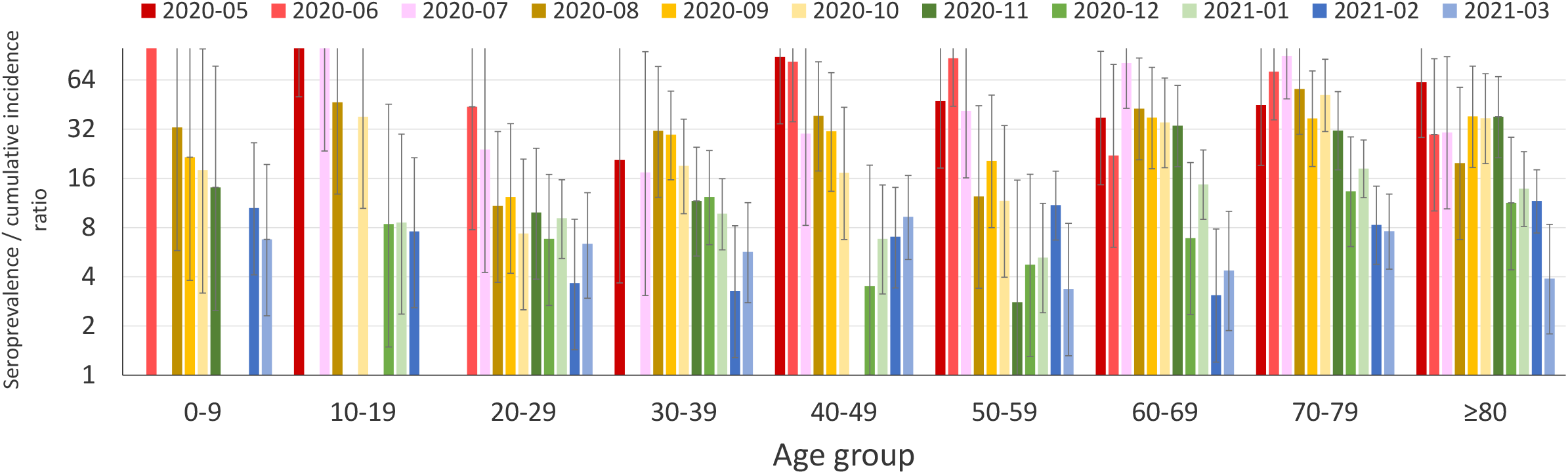
The ratio of seroprevalence to cumulative incidence in the Tokyo Metropolitan Area, May 2020–March 2021. The ratio of “SARS-CoV-2 seroprevalence at an indicated month” to “the cumulative incidence of reported COVID-19 from January 2020 to the indicated month” was calculated. The ratio corresponds to the actual number of infected cases per reported case. Data are blank for months when no samples were positive for SARS-CoV-2.

In the early phase of the pandemic, diagnostic tests detected as few as one case in more than 10 infections (i.e., an ascertainment rate of < 10%). The rate improved over time such that by March 2021 one case in about 3–10 infections was detected (i.e., an ascertainment rate of approximately 10–33%). The same trend was observed for all age groups, and the ascertainment rates did not differ substantially among the age groups.

The proportion of samples positive for the SARS-CoV-2 spike protein dramatically increased with the rollout of vaccination (i.e., after April 2021) (Figure 2). However, the seropositive proportions were slightly lower than the vaccination rates in people 70–79 and ≥ 80 years old. Furthermore, seropositive rates peaked and then declined for those aged 50 and over. The administration of the third vaccination after January 2022 restored the drop in seroprevalence (Figure 2A).

The SARS-CoV-2 seroprevalence in 0–9-year-olds increased during the Delta-dominant fifth and Omicron-dominant sixth epidemic waves, which started in July 2021 and January 2022, respectively (Figure 2A, B). Vaccination of children aged at least 5 years was approved and administered after February 2022 in Japan. Therefore, we subdivided the data for the 0–9 age group into 0–5 months old, 6 months–4 years old, and 5–9 years old (Figure 2C, D). The first subset age group showed a very high seroprevalence compared to the other two subset age groups. The seroprevalence for the two latter groups was low but increased after August 2021, reaching 8.0% for the 6 months–4 years and 9.3% for the 5–9 years groups in December 2021.

We also tested samples from Hokkaido, a prefecture situated about 800 km north of Tokyo. This prefecture reported an even higher number of cases per capita than the Tokyo Metropolitan Area during the first COVID-19 epidemic wave (Figure 1B); however, the second wave did not affect them much (Figure 1C).

A total of 17,079 serum samples from Hokkaido were collected and analyzed (Supplementary Table 2). The results were analogous to those obtained from the Tokyo Metropolitan Area (Figure 4). The seroprevalence was less than 5% for all age groups until the implementation of the vaccination program. The seroprevalence increased as the vaccines were administered, although the older age groups showed lower seropositivity rates compared with their vaccination rates. We could not subdivide and analyze the seroprevalence data of children less than 10 years old in Hokkaido because of the low number of samples for this age group (Supplementary Table 2).

**Figure 4.**
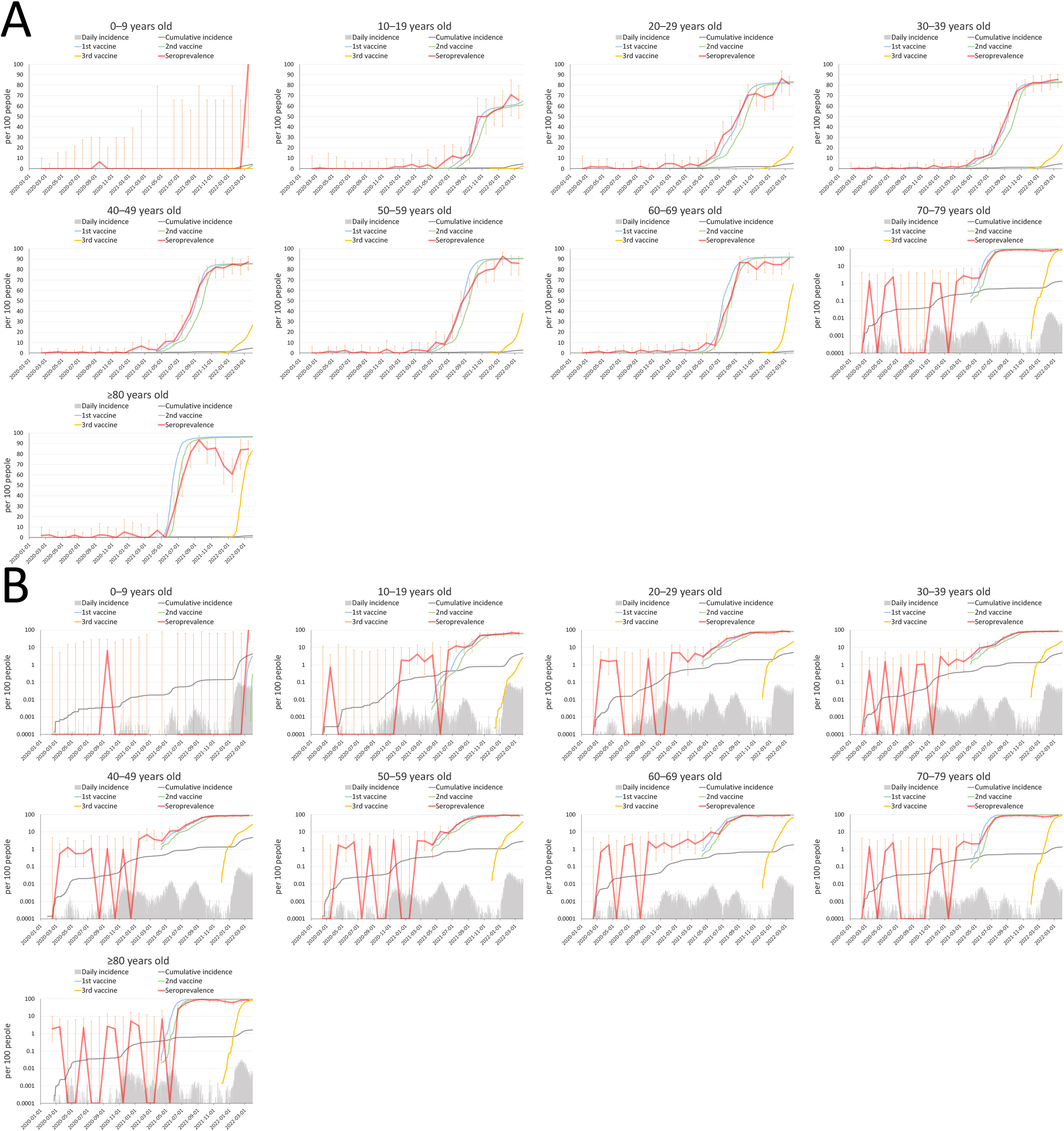
Seroprevalence of SARS-CoV-2 by age in Hokkaido, February 2020–March 2022. A) Seropositive rates for the SARS-CoV-2 spike protein in Hokkaido are shown by age group. The daily number of reported COVID-19 cases, their cumulative number, and the cumulative number for the first, second, and third vaccine administrations per population are also shown. The vertical orange lines indicate the 95% confidence intervals of the seroprevalence data. B) The same data on a logarithmic scale are shown.

## Discussion

The present study investigated the time course of seroprevalence of antibodies against SARS-CoV-2 by age group, analyzing over 60,000 samples from Japan. Although several previous studies have reported SARS-CoV-2 seroprevalence in Japan (18,19), our study is the largest to collect samples from a wide area over a period of 25 months. Diagnostic testing to identify SARS-CoV-2-infected people is important for gaining a better understanding of the epidemiological situation of COVID-19; however, it is virtually impossible to contain the epidemic through testing alone (20). The incidence and mortality rates of COVID-19 per population are considerably low in Japan (21). Yet, it has been suggested that the low number of tests per population caused many cases of infection to go undetected, and the reported statistics did not reflect the actual situation (22).

Our data show that approximately 5% of the Japanese population had been infected with SARS-CoV-2 by January 2021. That figure is much higher than the reported number of COVID-19 cases. Still, the low incidence rate was in stark contrast to other countries with a seroprevalence of > 30% at that time (3). Nonpharmaceutical interventions, such as physical distancing and wearing a face mask, play a significant role in controlling the COVID-19 pandemic, especially in the pre-vaccination era. Although Japan has not implemented a “lock-down,” the country issued a state of emergency—asking people to stay at home and limit mass gatherings and asking businesses, including restaurants and bars, to reduce their hours or close when COVID-19 cases surged (23). The country also implemented a unique strategy focusing on case-clusters (24,25).

Because we measured antibodies for the viral spike protein, we could not differentiate immunity by natural infection from immunity by vaccination after February 2021. Interestingly, the seroprevalence among children who were not yet eligible for vaccination in December 2021 was still as low as 10% in Japan. Thereafter, the infection was spread among children during the Omicron-dominant sixth epidemic wave (26), and their seropositive rates gradually increased at the beginning of 2022. The especially high seroprevalence among children aged 0–5 months after August 2021 must be the result of antibodies transferred from vaccinated mothers (Figure 2C, D).

The same low SARS-CoV-2 seroprevalence was observed in two far-apart regions: the Tokyo Metropolitan Area and Hokkaido. The first COVID-19 epidemic wave in Hokkaido preceded the other regions in Japan, and the second wave was well contained there (Figure 1). It was suggested that a large number of infections had been missed and that the Hokkaido population might have already attained herd immunity (27). Our results clearly show that this was not the case. Most of the population had not yet been infected with the virus.

Japan has achieved a high rate of SARS-CoV-2 seroprevalence among adults due to vaccinations since April 2021. In particular, most individuals in their 20s–40s, who were considered to play a major role in driving the COVID-19 epidemic (28), had been vaccinated by August 2021. After that, the country experienced an exceptionally low level of disease spread from September to November 2021 (Figure 1). However, it faced another wave after January 2022, possibly due to the introduction of the Omicron variant and the waning of vaccine-generated immunity.

A low seroconversion rate by vaccination and rapid immunity waning in the elderly have been reported at the individual level (29,30). In the present study, we observed this effect at the population level. In addition to vaccinating the elderly, who are at a high risk of developing severe illness, reducing their exposure to the virus should be key to protecting this vulnerable population. Booster shots also helped provide a high degree of population immunity.

In this study, we measured the antibody titers for the RBD of SARS-CoV-2 spike protein. Therefore, samples from both infected people and from vaccinated people showed positive results. Although the measurement of the antibody titers for the nucleoprotein can reflect only a history of natural infection with SARS-CoV-2, in our study, the sensitivity and specificity were not as high as the test for the spike protein (Supplementary Figure 1). The low sensitivity might have been due to the weak immunogenicity of the nucleoprotein, and the low specificity may be due to cross-reactivity between the seasonal coronavirus and SARS-CoV-2.

By testing antibodies for the spike protein, we discussed the actual incidence of COVID-19 in a pre-vaccination era. We validated the considerably high sensitivity and specificity of the test. Still, the estimate cannot be 100% accurate. Besides, our test subjects may not represent the general public in Japan. Our samples were from patients who visited healthcare facilities for various reasons other than COVID-19. People with underlying diseases could be more cautious about healthcare issues and avoid high-risk behavior, or patients with some symptoms could have had a high pretest probability of past infection with SARS-CoV-2. Other limitations include the small number of samples for the early phase of the study period, especially from children. Lastly, diagnostic histories of COVID-19 and vaccination status were not available for our analysis.

Our study highlights the very low SARS-CoV-2 infection rate in Japan. It also unveiled a hurdle to maintaining a high degree of population immunity among the elderly. In future studies, we should investigate how population immunity has affected and will affect the course of the pandemic. We must explore the levels of immunity required to prevent infection, hospitalization, and death from different SARS-CoV-2 variants. As the present study suggests that most populations in Japan have not yet been infected with the virus, the country’s current and future paths regarding the COVID-19 pandemic may continue to hold the world’s attention.

## Data Availability

All data produced in the present work are contained in the manuscript.

## Acknowledgments

We thank Tomoka Nagashima, Chiaki Kawana, Naoko Mizutani, Kyoko Yokota, Mikuru Sato, and Shihomi Kuwano at University of Tokyo; Noriaki Harada, Izuru Iwase, and Junya Shigenaga at Eiju General Hospital; Kazumi Yanagi at Japanese Red Cross Ashikaga Hospital; clinical laboratory staff at Keio University Hospital; Taro Kaneda, Ryoko Karakama, Yuka Suzuki, Shoma Ookaji, Ayano Aketa, Sayaka Sato, Misako Sugaya, Misa Hayasaka, Yoko Kira, Yuki Fujiwara, Akimi Kaku, and Masako Moteki at BML clinical laboratory staffs of National Hospital Organization Saitama Hospital; and Masami Isizaki at Zama Children’s Clinic for their efforts with the organization and collection of blood samples. We thank Kozue Amemiya, Kayako Chishima, Aya Fujiwara, Yoko Hamasaki, Naomi Ikeda, Tadatsugu Imamura, Takeaki Imamura, Keiya Inoue, Sachi Ishida, Mariko Kanamori, Tsuyoki Kawashima, Yura K Ko, Tomoe Mashiko, Rie Masuda, Reiko Miyahara, Konosuke Morimoto, Shohei Nagata, Yoshifumi Nin, Kota Ninomiya, Yukiyo Nitta, Hitoshi Oshitani, Mayuko Saito, Akiko Sakai, Eiichiro Sando, Kazuaki Sano, Asako Sato, Akiko Sayama, Yugo Shobugawa, Motoi Suzuki, Ayaka Takeuchi, Hiroto Tanaka, Fumie Tokuda, Naho Tsuchiya, Shogo Yaegashi, Yoko Yamagiwa, Lisa Yamasaki, Ikkoh Yasuda, and Fumi Yoshimatsu for the collection and management of data from public database and press releases. We also thank Susan Watson for editing the manuscript.

## Supplementary materials

**Supplementary Table 1.** Number of samples analyzed in the Tokyo Metropolitan Area.

|  |  | Age |  |  |  |  |  |  |  |  | Total |
| --- | --- | --- | --- | --- | --- | --- | --- | --- | --- | --- | --- |
|  |  | 0–9 | 10–19 | 20–29 | 30–39 | 40–49 | 50–59 | 60–69 | 70–79 | ≥ 80 |  |
| Month | 2020-02 | 6 | 6 | 32 | 41 | 67 | 97 | 120 | 182 | 135 | 686 |
|  | 2020-03 | 65 | 32 | 38 | 49 | 92 | 136 | 141 | 199 | 158 | 910 |
|  | 2020-04 | 79 | 48 | 20 | 33 | 30 | 34 | 50 | 45 | 27 | 366 |
|  | 2020-05 | 121 | 71 | 95 | 170 | 193 | 337 | 491 | 635 | 371 | 2484 |
|  | 2020-06 | 137 | 87 | 63 | 118 | 244 | 350 | 397 | 592 | 362 | 2350 |
|  | 2020-07 | 156 | 73 | 54 | 114 | 204 | 303 | 414 | 531 | 320 | 2169 |
|  | 2020-08 | 167 | 155 | 144 | 113 | 240 | 278 | 364 | 492 | 347 | 2300 |
|  | 2020-09 | 153 | 79 | 97 | 197 | 175 | 234 | 294 | 478 | 312 | 2019 |
|  | 2020-10 | 140 | 82 | 136 | 221 | 196 | 243 | 324 | 461 | 332 | 2135 |
|  | 2020-11 | 146 | 74 | 111 | 220 | 201 | 261 | 320 | 492 | 301 | 2126 |
|  | 2020-12 | 176 | 85 | 118 | 199 | 129 | 207 | 280 | 378 | 242 | 1814 |
|  | 2021-01 | 148 | 81 | 134 | 244 | 213 | 298 | 358 | 554 | 355 | 2385 |
|  | 2021-02 | 181 | 103 | 99 | 164 | 186 | 268 | 327 | 457 | 348 | 2133 |
|  | 2021-03 | 188 | 117 | 80 | 155 | 186 | 216 | 265 | 480 | 302 | 1989 |
|  | 2021-04 | 106 | 52 | 77 | 133 | 135 | 170 | 235 | 328 | 221 | 1457 |
|  | 2021-05 | 108 | 52 | 84 | 145 | 162 | 241 | 289 | 458 | 313 | 1852 |
|  | 2021-06 | 179 | 64 | 75 | 136 | 131 | 181 | 235 | 396 | 222 | 1619 |
|  | 2021-07 | 243 | 80 | 75 | 131 | 123 | 181 | 223 | 313 | 235 | 1604 |
|  | 2021-08 | 217 | 106 | 70 | 134 | 169 | 253 | 268 | 347 | 240 | 1804 |
|  | 2021-09 | 96 | 45 | 76 | 129 | 137 | 183 | 213 | 328 | 219 | 1426 |
|  | 2021-10 | 231 | 80 | 80 | 140 | 149 | 186 | 235 | 315 | 206 | 1622 |
|  | 2021-11 | 206 | 67 | 76 | 129 | 123 | 188 | 226 | 317 | 225 | 1557 |
|  | 2021-12 | 177 | 66 | 74 | 128 | 116 | 163 | 247 | 342 | 223 | 1536 |
|  | 2022-01 | 160 | 59 | 76 | 137 | 147 | 187 | 206 | 322 | 216 | 1510 |
|  | 2022-02 | 128 | 49 | 87 | 110 | 118 | 175 | 233 | 322 | 245 | 1467 |
|  | 2022-03 | 53 | 32 | 89 | 135 | 120 | 164 | 206 | 342 | 211 | 1352 |
|  | Total | 3767 | 1845 | 2160 | 3625 | 3986 | 5534 | 6961 | 10106 | 6688 | 44672 |

**Supplementary Table 2.** Number of samples analyzed in Hokkaido.

|  |  | Age |  |  |  |  |  |  |  |  | Total |
| --- | --- | --- | --- | --- | --- | --- | --- | --- | --- | --- | --- |
|  |  | 0–9 | 10–19 | 20–29 | 30–39 | 40–49 | 50–59 | 60–69 | 70–79 | ≥ 80 |  |
| Month | 2020-02 | 34 | 28 | 28 | 65 | 79 | 58 | 79 | 82 | 53 | 506 |
|  | 2020-03 | 72 | 140 | 155 | 213 | 361 | 214 | 288 | 285 | 121 | 1849 |
|  | 2020-04 | 21 | 22 | 64 | 147 | 155 | 116 | 113 | 120 | 68 | 826 |
|  | 2020-05 | 20 | 30 | 56 | 130 | 181 | 89 | 116 | 137 | 58 | 817 |
|  | 2020-06 | 14 | 49 | 66 | 151 | 174 | 115 | 130 | 125 | 89 | 913 |
|  | 2020-07 | 10 | 62 | 76 | 141 | 89 | 95 | 95 | 121 | 71 | 760 |
|  | 2020-08 | 9 | 60 | 48 | 95 | 67 | 68 | 72 | 78 | 39 | 536 |
|  | 2020-09 | 15 | 50 | 88 | 98 | 93 | 57 | 71 | 88 | 38 | 598 |
|  | 2020-10 | 15 | 51 | 78 | 89 | 93 | 69 | 85 | 89 | 53 | 622 |
|  | 2020-11 | 9 | 34 | 68 | 88 | 104 | 56 | 85 | 93 | 40 | 577 |
|  | 2020-12 | 9 | 54 | 61 | 97 | 81 | 55 | 76 | 102 | 38 | 573 |
|  | 2021-01 | 6 | 57 | 62 | 108 | 82 | 67 | 94 | 89 | 36 | 601 |
|  | 2021-02 | 3 | 49 | 69 | 95 | 73 | 65 | 79 | 81 | 26 | 540 |
|  | 2021-03 | 0 | 65 | 87 | 130 | 55 | 68 | 99 | 107 | 35 | 646 |
|  | 2021-04 | 1 | 27 | 67 | 120 | 99 | 56 | 85 | 99 | 29 | 583 |
|  | 2021-05 | 0 | 32 | 54 | 85 | 100 | 58 | 82 | 97 | 47 | 555 |
|  | 2021-06 | 2 | 29 | 58 | 97 | 103 | 61 | 82 | 101 | 31 | 564 |
|  | 2021-07 | 2 | 57 | 56 | 91 | 62 | 51 | 88 | 96 | 32 | 535 |
|  | 2021-08 | 3 | 60 | 68 | 120 | 96 | 64 | 105 | 105 | 43 | 664 |
|  | 2021-09 | 1 | 29 | 63 | 106 | 88 | 58 | 75 | 84 | 45 | 549 |
|  | 2021-10 | 2 | 26 | 54 | 116 | 101 | 64 | 75 | 90 | 45 | 573 |
|  | 2021-11 | 2 | 30 | 64 | 109 | 73 | 63 | 81 | 102 | 28 | 552 |
|  | 2021-12 | 2 | 47 | 66 | 97 | 92 | 57 | 64 | 97 | 32 | 554 |
|  | 2022-01 | 1 | 29 | 48 | 97 | 81 | 69 | 79 | 69 | 33 | 506 |
|  | 2022-02 | 2 | 24 | 44 | 90 | 99 | 52 | 79 | 94 | 25 | 509 |
|  | 2022-03 | 1 | 32 | 74 | 117 | 95 | 57 | 65 | 91 | 39 | 571 |
|  | Total | 256 | 1173 | 1722 | 2892 | 2776 | 1902 | 2442 | 2722 | 1194 | 17079 |

**Supplementary Figure 1.**
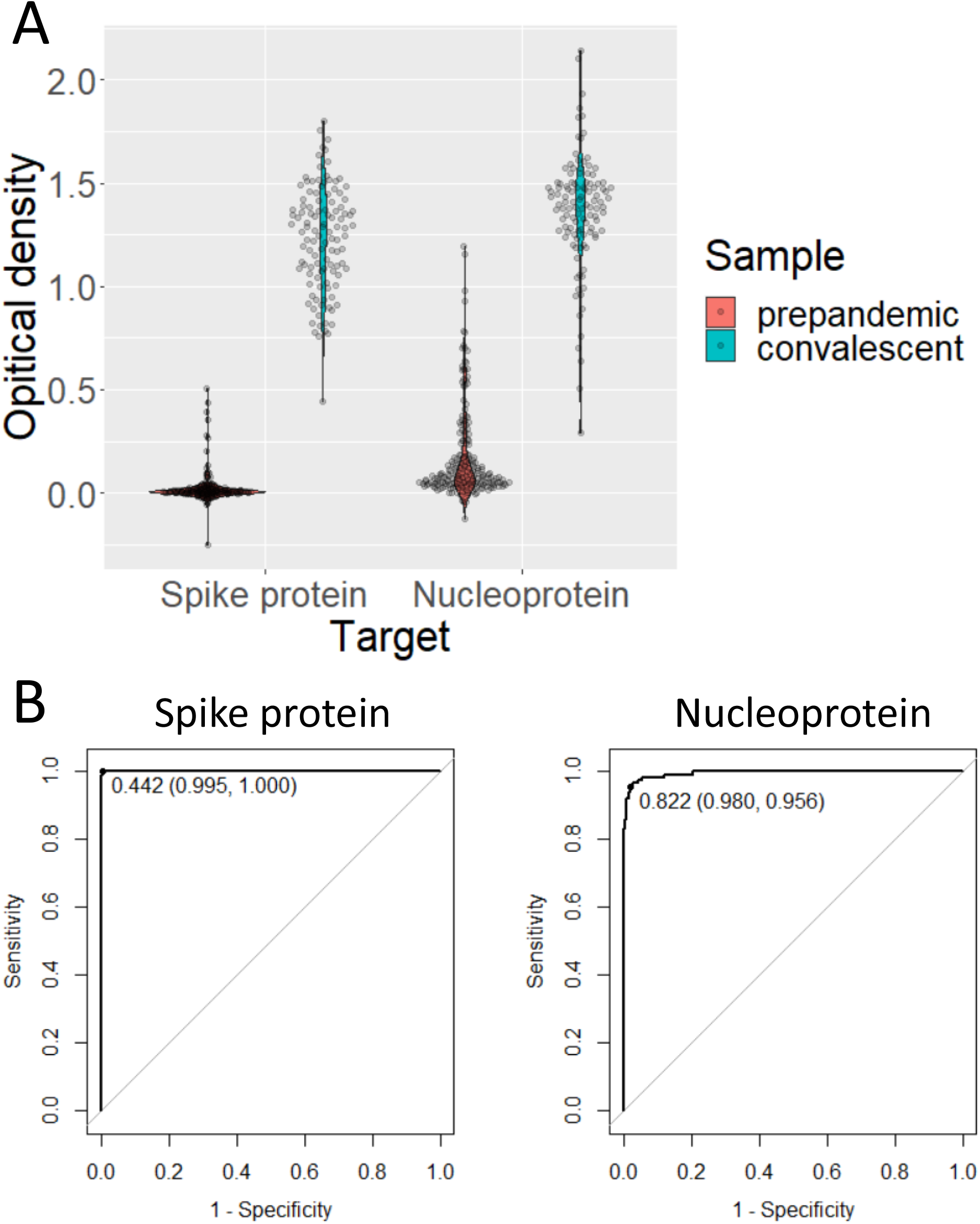
Antibody titers for SARS-CoV-2 in prepandemic and convalescent samples. A) The results of ELISA tests against the SARS-CoV-2 spike protein and nucleoprotein are shown for samples collected in 2012 (prepandemic, n = 200) and samples from COVID-19-confirmed cases (convalescent, n = 113). B) ROCs were drawn for the SARS-CoV-2 spike protein and nucleoprotein to differentiate prepandemic and convalescent samples. A cut-off value was determined by using Youden’s index. The specificity and sensitivity of the test are described in parentheses in the figure.

